# TRIDENT: a framework for robust multi-trait GWAS identifies 66 novel multi-trait osteoarthritis signals

**DOI:** 10.64898/2026.08.12.26360256

**Authors:** Yahong Wu, Sirine Saafi, Shibo Chen, Ziyi Xiong, Mijin Jung, Lorraine Southam, Genetics of Osteoarthritis Consortium, Benjamin G. Faber, Manfred Kayser, Joyce BJ van Meurs, Eleftheria Zeggini, Cindy G. Boer

## Abstract

As multi-trait genome-wide association studies (GWAS) are increasingly used to identify shared genetic associations across related phenotypes, practical approaches to assess the robustness of their findings are lacking. Here we present a three-step framework (Trident) for robust multi-trait GWAS that uses an earlier, smaller GWAS meta-analysis to test whether phenotypes can be validly combined as well as the latest, largest GWAS meta-analysis of the same phenotypes for discovery, followed by translational annotation to assess disease relevance and prioritize likely effector genes. We applied Trident by using the Combined-GWAS (C-GWAS) method to osteoarthritis, a degenerative joint disease, across five osteoarthritis joint sites. Signals identified in the earlier GWAS meta-analysis showed high validation in the replication dataset, supporting the robustness of this approach. Applied to the latest and largest osteoarthritis GWAS meta-analysis, C-GWAS identified 66 novel associations not identified with conventional single-trait GWAS meta-analyses, including signals with shared and discordant effects across different joint sites. Translational annotation linked these signals to biologically plausible osteoarthritis genes and pathways. Together, we provide a practical framework for robust multi-trait GWAS that increases detection power by identifying novel signals and, by applying it to the example of osteoarthritis of five joints, refine the genetic architecture of this common disease.

## Introduction

Most common complex traits and diseases are influenced by both genetic and environmental factors, for which genome-wide association studies (GWAS) have been widely used, and were successful in unveiling their genetic architecture[1]. Over the past two decades, GWAS has evolved from small, single-cohort studies to large international multi-cohort studies comprising hundreds of thousands to millions of individuals[2, 3]. As a result of GWAS sample size expansions, GWAS databases such as the NHGRI–EBI GWAS Catalogue now hosts tens of thousands of published genetic associations across diverse traits, providing a comprehensive resource for secondary and integrative analyses[4]. Many common complex diseases are aetiologically heterogeneous and manifest as multiple related phenotypes that may share underlying biological pathways. Osteoarthritis exemplifies this complexity because it affects anatomically distinct joints, including the hip, knee, spine, finger and thumb. Although these joint-specific phenotypes are conventionally analysed in separate GWAS, previous studies have shown that they share part of their genetic basis[2, 3]. Multi-trait GWAS methods, which jointly analyse summary statistics from related phenotypes, capture shared genetic effects to identify novel variants that do not reach significance in individual single-trait analyses[5–7]. These multi-trait GWAS analyses have been increasingly applied to correlated traits of the same complex phenotype, or to traits with shared biological pathways[6, 8–11]. Yet despite the growing use, there remains no clear framework for assessing the robustness of the findings obtained in multi-trait GWAS.

In single-trait GWAS, robustness is typically established through replication in independent cohorts and, increasingly, through meta-analysis across multi-cohort studies [12]. This paradigm is difficult to apply directly to multi-trait GWAS, as they usually rely on the largest available GWAS meta-analysis summary statistics dataset for each phenotype and leave no comparable independent datasets for replication purposes. Moreover, novel signals identified through multi-trait GWAS are, by definition, not significant in the corresponding single-trait GWASs and may therefore remain difficult to replicate even when additional cohort data are available, particularly because such cohorts are often smaller than, or already included in, the meta-analyses dataset used for discovery. As a result, the traditional replication and meta-analysis framework cannot be directly transferred to multi-trait GWAS. Existing multi-trait methods, such as C-GWAS[5, 6] fastASSET[13] or MTAG[7], are validated during their development phase using simulations or selected example traits. However, when applied to a new set of input data, these tools are only expected to perform well if certain assumptions are met, both biologically and technically. Biologically, the input traits should share some degree of genetic basis; technically, each method may impose additional tool-specific assumptions. For example, MTAG assumes that genetic effects across input traits follow a shared variance–covariance structure[7], an assumption that may not hold for all phenotype combinations. If such conditions are violated, multi-trait GWAS may yield false-positive findings, and, if independent replication or comparable meta-analysis are unavailable, the overall results become harder to validate. Together, these challenges highlight the lack of a framework to assess the robustness of multi-trait GWAS.

To address these challenges, we present Trident, a practical analytical framework to support robust multi-trait GWAS findings while preserving discovery power. Trident relies on the availability of multiple GWAS datasets for the same multiple phenotypes, using the largest and most recent GWAS for discovery and earlier versions to assess whether the phenotype combination can be robustly analysed by the multi-trait tool. Trident comprises three steps (**Figure 1**). First, multi-trait GWAS is applied to an earlier and smaller dataset for the target phenotypes. The signals identified are evaluated in the latest and largest GWAS of the same phenotypes to assess whether they are reproducible in a higher-powered setting. Second, the same multi-trait GWAS approach is applied to the latest and largest GWAS datasets to maximize discovery. Third, the resulting associations are interpreted and further validated using multiple external lines of evidence, including quantitative trait loci, functional genomic annotations, and gene databases. Together, Trident provide a practical strategy for evaluating and interpreting multi-trait GWAS findings.

We applied this Trident to the example of osteoarthritis, a common and genetically complex disease affecting more than 500 million individuals worldwide[14]. Osteoarthritis is a particularly suitable setting for multi-trait GWAS because it affects multiple anatomically distinct joints, including the hip, knee, spine, finger, and thumb[15], which represent related manifestations of the same disease and are therefore expected to share at least part of their genetic basis[16]. We selected C-GWAS[5, 6] as the multi-trait method of choice for this study because it makes fewer restrictive assumptions about the shared genetic architecture of the input traits. Moreover, C-GWAS has demonstrated robust performance when analysing a large number of traits simultaneously, including highly correlated traits, such as over hundred ear morphology traits[17] and almost one thousand facial shape traits[6]. In addition, C-GWAS can effectively model heterogeneous effect patterns across traits, for example variants with opposite directions of effect across phenotypes, making it well suited to osteoarthritis genetics. Using this Trident, we analysed osteoarthritis at five joint sites with two generations of GWAS summary statistics from the Genetics of Osteoarthritis Consortium, using the earlier, smaller dataset of more than eight hundred thousand individuals (GO-1[2]) for validation and the latest, largest dataset of nearly two million individuals (GO-2[3]) for discovery.

## Results

To enable robust multi-trait GWAS, we developed and implemented a three-step analytical framework (Trident) consisting of (i) applying multi-trait GWAS to an earlier and smaller GWAS meta-analysis dataset for the target phenotypes, and evaluating the resulting signals in the latest and largest available GWAS meta-analysis of the same phenotypes, (ii) applying the same multi-trait GWAS approach to the latest and largest GWAS dataset to maximize discovery, and (iii) interpreting the resulting genetic associations through integration of external evidence (**Figure 1**). Applying Trident to osteoarthritis across five joint sites, we first analyzed the validation GWAS (GO-1[2] N=826,690) using C-GWAS. We then evaluated the resulting signals in the discovery dataset (GO-2[3], N =1,962,069) to assess robustness in a higher-powered setting. Next, we applied C-GWAS to the latest and largest GO-2 dataset to maximize genetic discovery. Finally, we integrated 22 independent lines of translational evidence to prioritize likely effector genes underlying the identified associations.

### Step 1: C-GWAS shows high validity in large-scale osteoarthritis GWAS

Although the multi-trait GWAS tool we chose, C-GWAS, has been validated in its development phase, the method was initially developed in the context of facial morphology using GWAS data from a relatively small sample (n = 10,115)[5]. To assess its performance for osteoarthritis across five joints (hip, knee, spine, finger, and thumb) in much larger datasets, we first applied C-GWAS to GWAS meta-analysis summary statistics from the earlier and smaller GO-1 dataset [2]. These five joint-specific osteoarthritis phenotypes were selected because they represent manifestations of the osteoarthritis at different site and are therefore expected to share at least part of their genetic basis, making them suitable for multi-trait analysis. Using GO-1 as input, C-GWAS identified 70 independent genome-wide significant (P ≤ 5 × 10⁻^8^) signals (**Supplementary Table 1**).

Then, we evaluated these 70 signals identified by assessing their true-positive and false-negative rates. The rationale is that signals detected in the earlier and smaller GWAS meta-analysis dataset (GO-1) are more likely to be genuine if they are additionally supported in the latest and largest GWAS meta-analysis dataset (GO-2) of the same phenotypes, where statistical power is larger. Overall, C-GWAS achieved a true-positive rate of 97.1% (68/70) across osteoarthritis traits. This consisted of 59 signals that had already been identified in at least one of the 11 osteoarthritis phenotype GWAS meta-analyses in GO-1, together with 9 of the 11 additional signals that were novel to GO-1 but replicated in GO-2 (**Figure 2a**). Of these 11 GO-1-novel signals, nine replicated at the Bonferroni-corrected 0.05 significance threshold (P ≤ 4.13 × 10⁻⁴), and six of those reached genome-wide significance (P ≤ 5 × 10⁻⁸) in at least one of the 11 osteoarthritis phenotype GWAS meta-analyses in GO-2 **(Supplementary Table 2)**. The two remaining signals did not reach Bonferroni-corrected significance in any GO-2 phenotype: rs191515286 (minimal p value across GWAS (minP) = 0.01, minor allele frequency (MAF) = 0.2%) and rs191129345 (minP = 0.06, MAF = 0.1%). However, both were located within osteoarthritis-associated loci identified in GO-2, though not in linkage disequilibrium (LD, r^2^<0.1) with the reported lead variants[3].

Next, we assessed the false-negative rate of C-GWAS by examining its ability to rediscover signals identified by GO-1. In the original GO-1 study, 71 independent signals were identified across five osteoarthritis phenotypes (hip, knee, spine, finger, and thumb osteoarthritis), of which 69 were included in the C-GWAS analysis (two removed due to quality control (**Methods**)). Among these 69 signals, 56 (81.2%) were re-identified at genome-wide significance (P < 5 × 10⁻⁸), and all 69 (100%) were re-identified at genome-wide suggestive significance (P < 1 × 10⁻⁶) in the C-GWAS results (**Supplementary Table 3**).

Together, these results support the empirical robustness of C-GWAS in large-scale multi-trait osteoarthritis GWAS meta-analysis, with 100% recovery of established associations at genome-wide suggestive significance, a true-positive rate of 97.1%, and identifying 11 additional loci representing a 15.5% increase in discovery power.

### Step 2: C-GWAS identifies 66 novel genetic osteoarthritis signals

As the second step, we aimed to identify novel multi-trait genetic signals for osteoarthritis by applying C-GWAS to the most recent and largest osteoarthritis GWAS dataset, the GO-2 meta-analysis summary statistics[3](n=1,962,069), which is twice as large as the earlier GO-1 dataset[2] (n = 826,690). As input for C-GWAS, we used the same five GWAS meta-analysis summary statistics for osteoarthritis joint sites (hip, knee, spine, finger, and thumb) from GO-2. As a result, C-GWAS identified 297 independent signals from 169 genetic loci (**Table 1, Supplementary Table 4, Figure 2**).

**Table 1.**
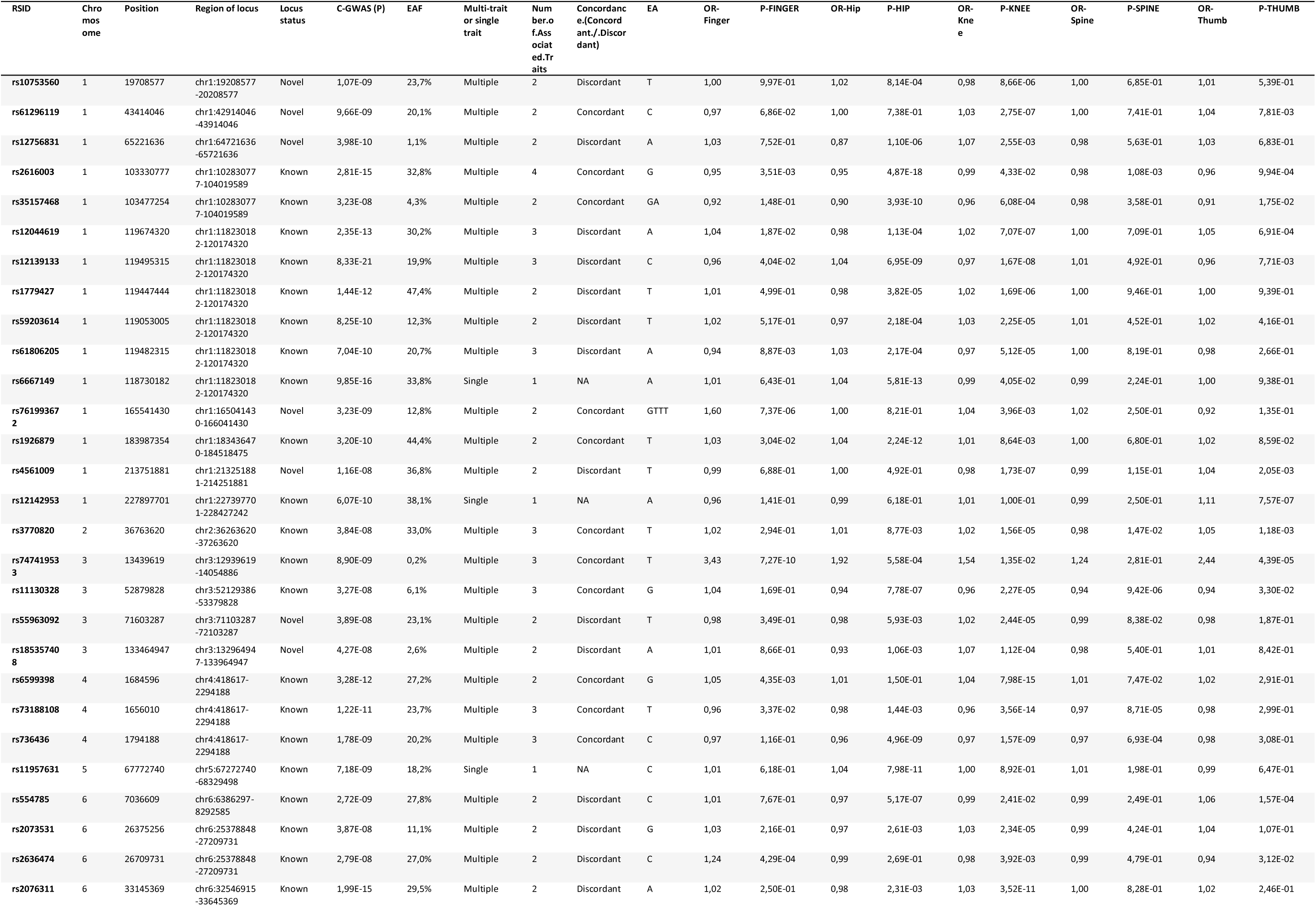
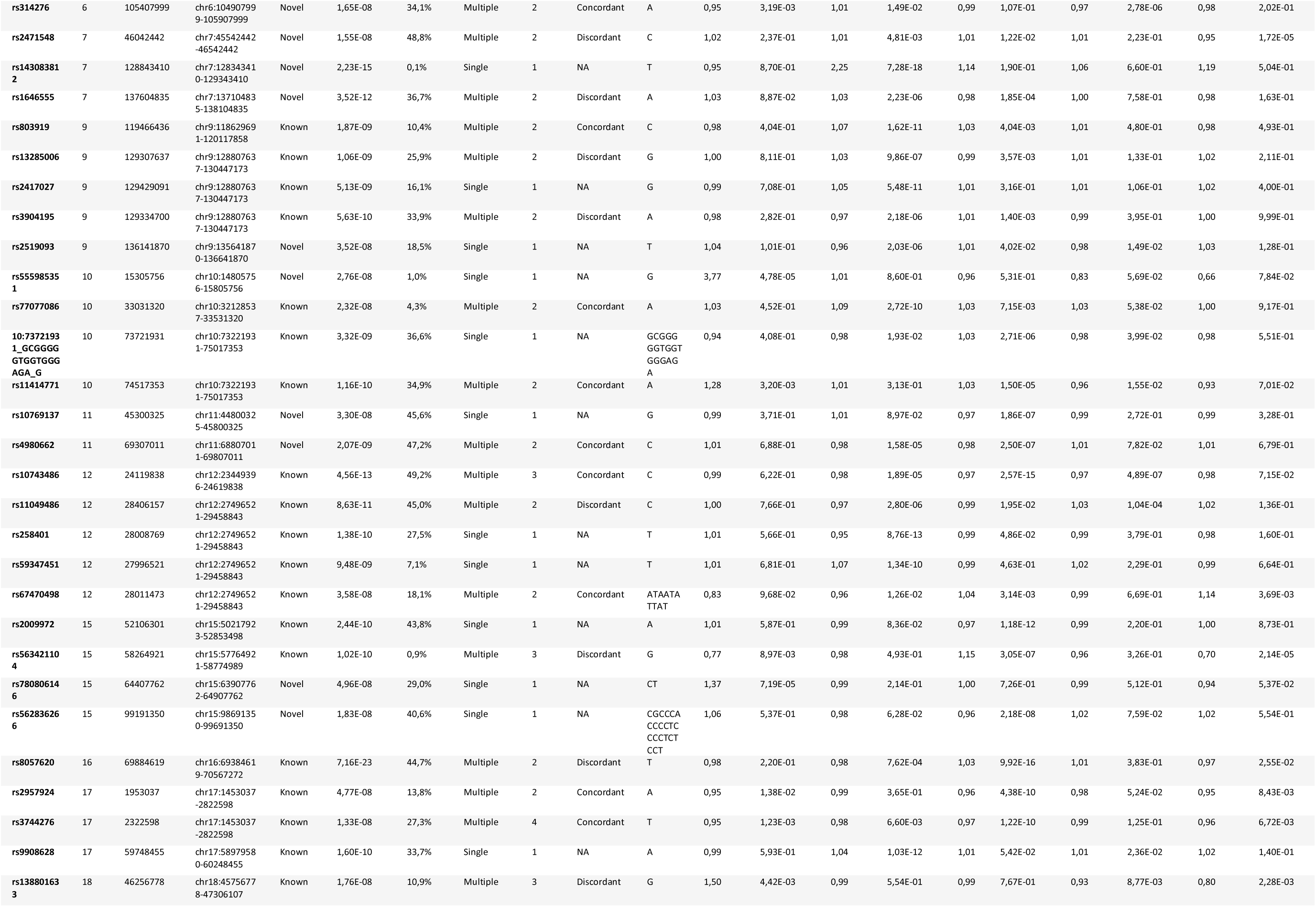
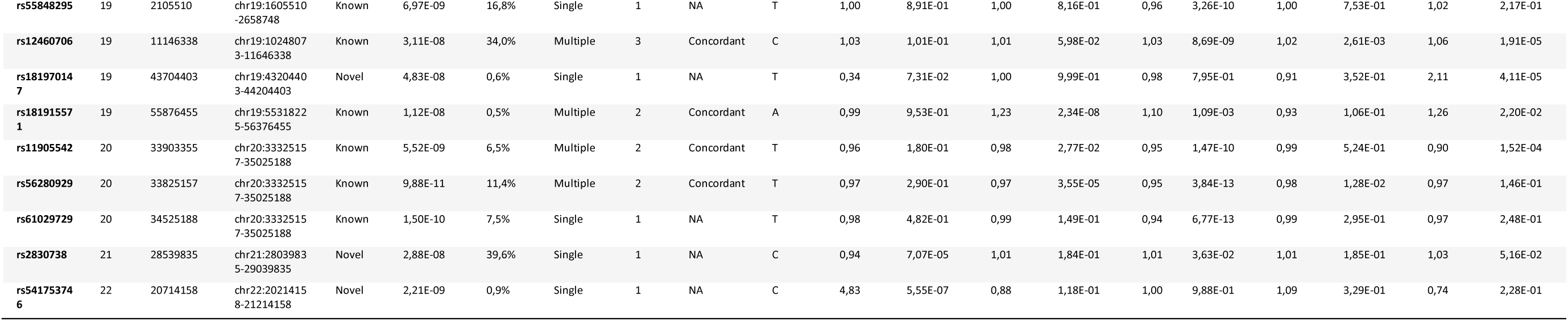
Novel osteoarthritis association signals identified by C-GWAS. The table presents 66 independent genome-wide significant osteoarthritis association signals identified by C-GWAS (P ≤ 5 × 10⁻^8^). Of these, 20 were located in novel osteoarthritis loci, and 46 represented additional independent signals within known osteoarthritis loci. Locus status indicates whether a signal was located within a locus previously reported for osteoarthritis in GO-2.Chromosome, position and locus region are reported using the hg19 reference genome. RSID denotes the dbSNP identifier; a chromosome-position-allele identifier is provided where an RSID was unavailable. C-GWAS (P) value is the association (P) value from the C-GWAS analysis, and EAF denotes the frequency of the effect allele. The multi-trait or single-trait classification indicates whether a signal was associated with multiple joint-specific phenotypes or a single joint-specific phenotype, according to the definition provided in the Methods. Concordance indicates whether the directions of effect were consistent across the associated joint-specific phenotypes; this classification is not applicable to single-trait signals. Joint-specific odds ratios and (P) values from the five GO-2 GWAS used as inputs to C-GWAS are also reported.Abbreviations: C-GWAS, combined genome-wide association study; EA, effect allele; EAF, effect allele frequency; GO-2, Genetics of Osteoarthritis consortium study 2; OA, osteoarthritis; OR, odds ratio; RSID, reference SNP cluster identifier.

**Table 2.**
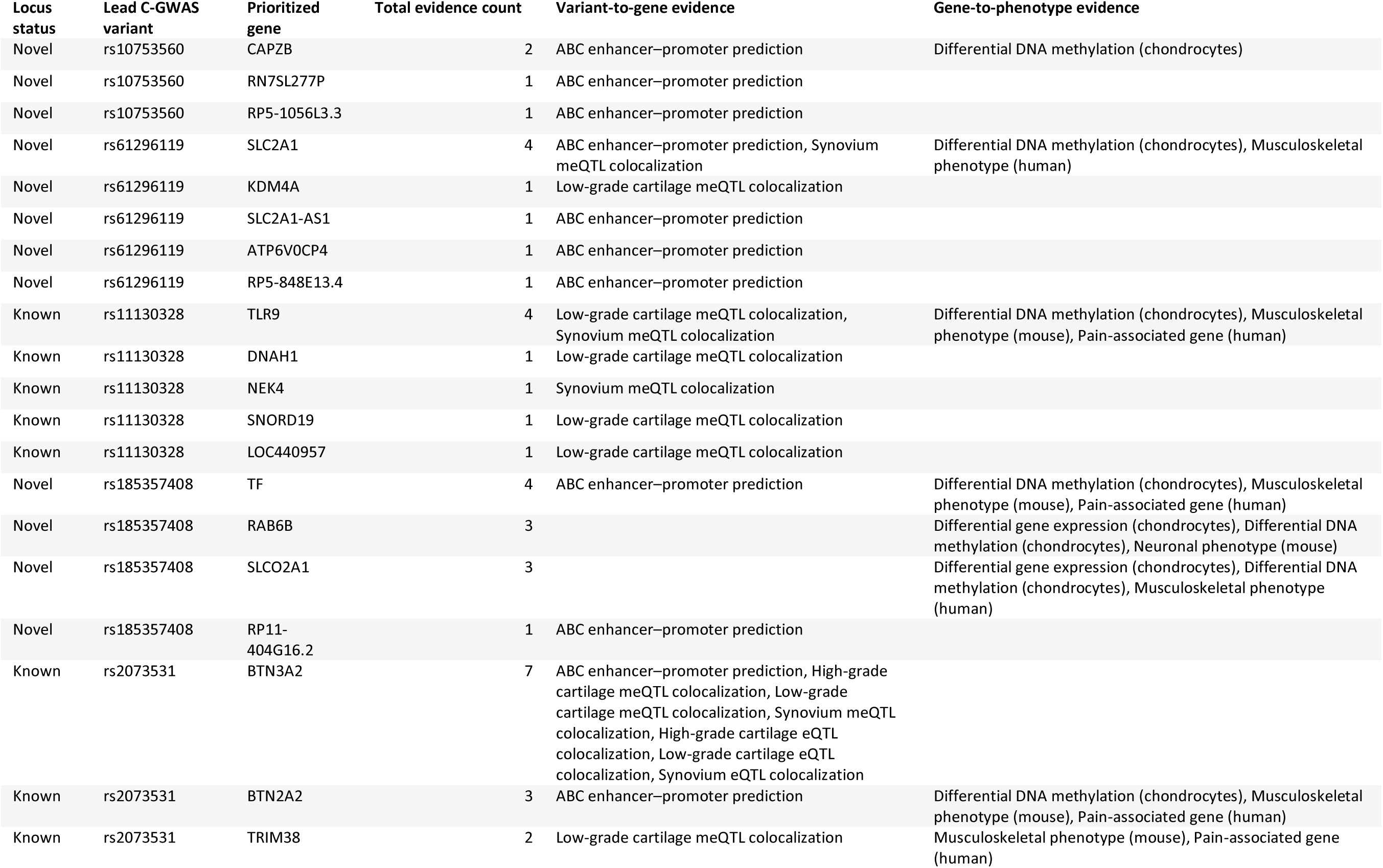
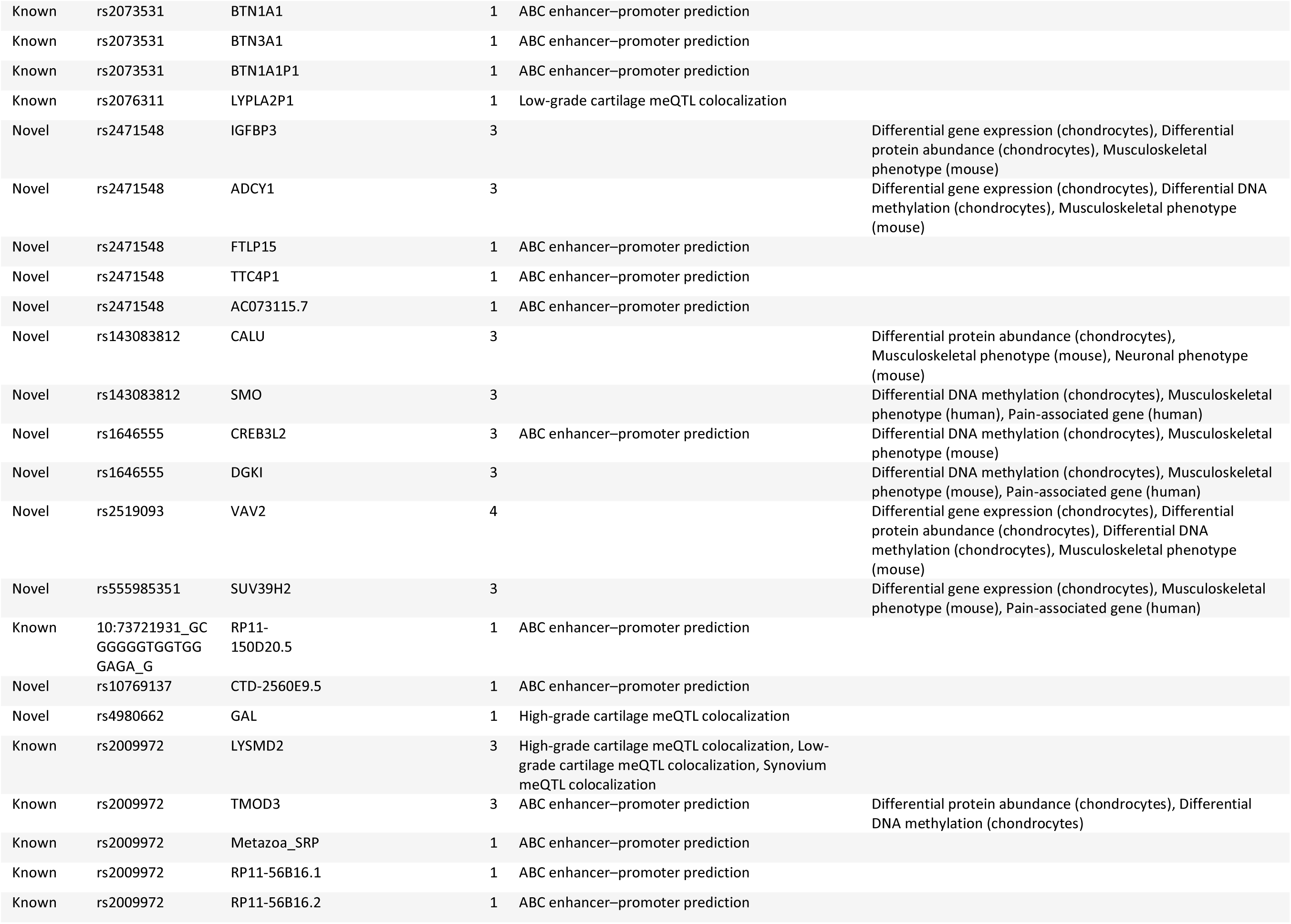
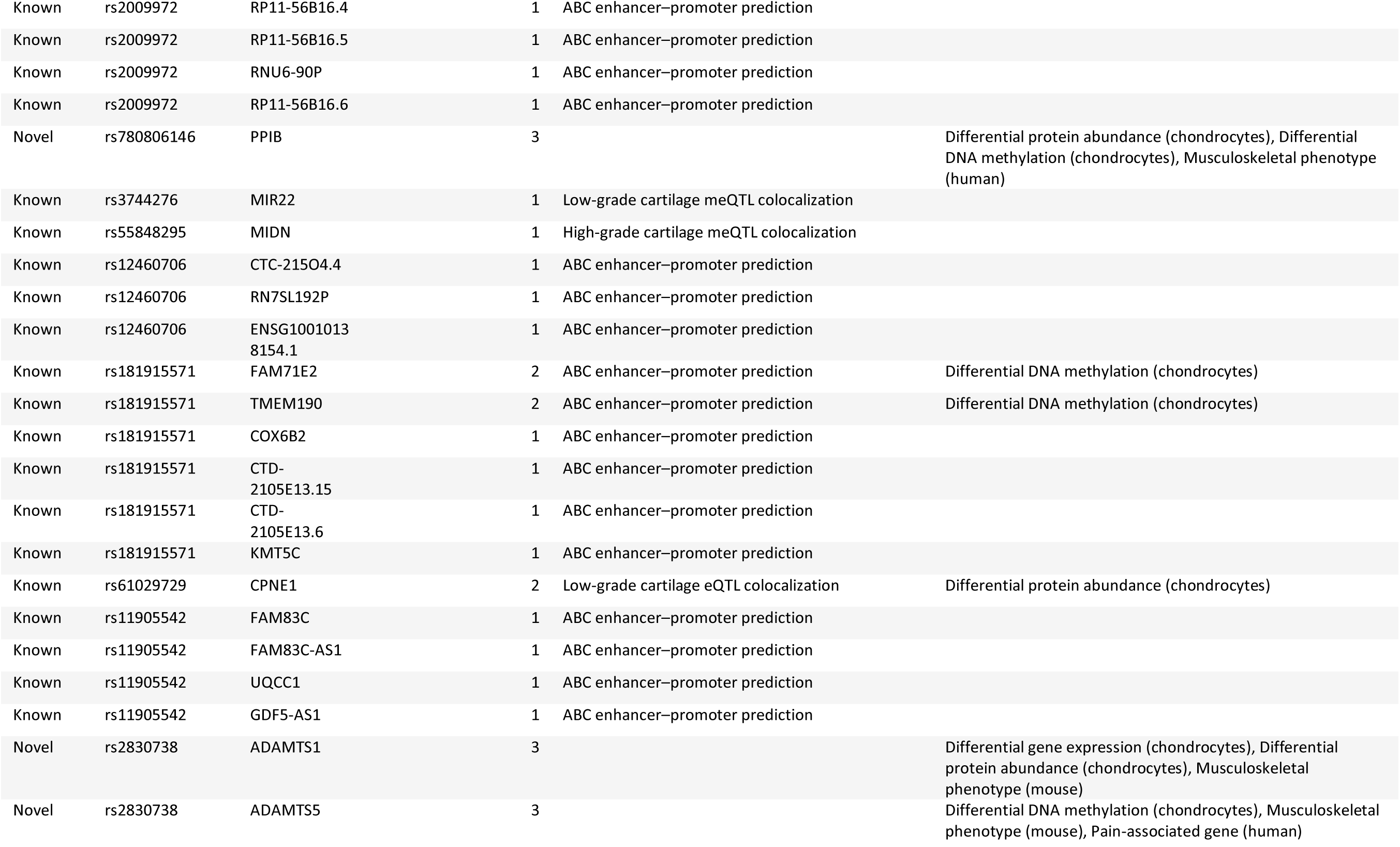
Prioritized likely effector genes at loci containing novel C-GWAS osteoarthritis association signals. The table presents 66 likely effector genes prioritized at loci containing the novel C-GWAS association signals reported in Table 1. Of these, 28 genes were prioritized at 12 novel osteoarthritis loci, and 38 were prioritized at 10 loci previously reported for osteoarthritis in GO-2. Locus status indicates whether the locus was novel or previously reported in GO-2. Genes were prioritized by integrating variant-to-gene and gene-to-phenotype evidence, as described in the Methods. A gene was included when it was supported by at least one significant variant-to-gene evidence source or at least three gene-to-phenotype evidence sources. Total evidence count indicates the total number of supporting evidence types identified for each gene. Multiple evidence types within the same category are separated by commas. An empty cell indicates that no evidence of the corresponding category was identified. Variant-to-gene evidence included activity-by-contact enhancer–promoter predictions and colocalization with methylation, expression or protein quantitative trait loci in osteoarthritis-relevant tissues. Gene-to-phenotype evidence included chondrocyte molecular phenotypes and relevant mouse and human phenotype annotations. The specific variants, molecular traits, tissues, CpG sites and directions of effect underlying each evidence type are reported in Supplementary Table 9. Gene prioritization identifies candidates for functional investigation but does not establish a causal relationship between a gene and osteoarthritis. Abbreviations: ABC, activity-by-contact; C-GWAS, combined genome-wide association study; eQTL, expression quantitative trait locus; GO-2, Genetics of Osteoarthritis consortium study 2; meQTL, methylation quantitative trait locus; OA, osteoarthritis; pQTL, protein quantitative trait locus.

To identify additional osteoarthritis associations, we compared the 297 C-GWAS signals with the complete reference catalogue of 339 independent signals from GO-2 across all 11 osteoarthritis phenotypes. Of the 297 C-GWAS signals, 231 were already identified in the GO-2 and the remaining 66 signals were novel osteoarthritis signals (**Table 1**), representing a 19.5% increase (66/339) in discovery power. These 66 signals, which is the principal discovery of this analysis, included 20 signals in 20 previously unidentified loci and 46 conditionally independent secondary signals across 28 known loci.

In these 28 known loci, all primary signals identified by GO-2 were rediscovered at genome-wide significance (P < 5 × 10⁻8) in the C-GWAS results (**Supplementary Table 4**), indicating that these 46 novel signals represented independent, secondary associations at known loci. Among these 46 secondary signals in known loci, 41 (89.1%) had MAF ≥5%. Similarly, among the 20 signals in novel loci, 14 (70%) had MAF ≥5%, with only three having MAF <1% (**Figure 3**). Those observations indicated that the C-GWAS result was not disproportionately driven by rare variants.

For benchmarking of the discovery gain by C-GWAS, we compared C-GWAS results to specific GO-2 analyses results (**Supplementary Table 5, Figure 2**) [3]. Compared to the 308 signals GO-2 identified from the same the five osteoarthritis phenotypes, C-GWAS detected 70 additional signals, corresponding to a 22.7% increase in discovery power. Compared to the 175 signals identified from the single “All OA” phenotype, representing the largest conventional single-trait GWAS in osteoarthritis (n= 1,962,069), C-GWAS identified 158 additional signals, representing a 90.3% increase in discovery power. These results show that, beyond increasing sample size as in traditional GWAS meta-analysis, using shared genetic architecture across traits through multi-trait GWAS can further increase genetic discovery.

### Multi-trait and discordant genetic effects across osteoarthritis joints

A defining feature of multi-trait analysis by C-GWAS is the ability to detect variants influencing multiple related traits, regardless of effect direction. So, we next systematically characterized the multi-trait signals in osteoarthritic joints, defined as significant association with at least two osteoarthritis joints (**Methods**). As a result, of the total 297 genome-wide significant signals highlighted by C-GWAS, 220 (74.1%) demonstrated multi-trait osteoarthritis effects (**Supplementary Table 4, Figure 2**, **Figure 4**). Five signals were associated with all five joint sites, and 18 were associated with four joint sites (**Figure 4**). Among the multi-trait 220 signals, 46 were novel signals discovered in this study (11 from novel loci and 35 from known loci) (**Supplementary Table 4**).

Notably, 42 of the 220 multi-joint signals exhibited discordant effects across different joints (**Supplementary Table 4, Figure 4B**), with the same allele increasing risk at one joint while decreasing risk at another one. Among these 42 multi-trait discordant signals, one discordant signal was associated with all five joints, and three were with four. To further evaluate the robustness of these discordant associations, we performed a sensitivity analysis using osteoarthritis GWAS summary statistics from the Rotterdam Study (RS), which are based exclusively on radiographically defined osteoarthritis[3]. Because radiographic osteoarthritis assessment is standardized [15], this analysis allowed us to test whether the observed discordant effects were artefacts arising from mixed osteoarthritis definitions, such as self-reported questionnaires, used in some GO-2 cohorts[3], rather than true biological differences. Of the 42 discordant signals identified in GO-2, 19 signals across 14 loci replicated in the RS GWAS, and showed the same discordant directions of effect on osteoarthritis in different joints (**Supplementary Table 6**).

To investigate patterns of discordance for a trait perspective, we examined each pair of joints (i.e., knee and hip) by the proportion of discordant effect variants in all multi-trait variants (**Figure 4C**). The highest rate of discordance was observed between the thumb and the hip, in which 22 signals were identified with an effect on both of these joints, while 30% of them had discordant effects. In contrast, thumb and finger osteoarthritis (N=70) had the highest agreement rate, where only 14% of shared variants had discordant effects.

### Step 3: Translational prioritization identifies 66 novel likely effector osteoarthritis genes

As the third step, we integrated multiple external lines of evidence to assess whether the identified association signals were truly linked to osteoarthritis and to prioritize their likely effector genes. Consistent with the GO-2 study[3], we used 22 “lines of evidence” categorized into variant-to-gene evidence (e.g., Activity-By-Contact (ABC) Model predictions, and molecular QTL colocalization[18, 19]) as well as gene-to-phenotype evidence (e.g., differential gene expression in osteoarthritis-related tissue[18–21], human and mouse mutational osteoarthritis related phenotypes), with full details provided in the **Methods** and **Supplementary Methods**.

We first focused on the 20 novel loci identified by comparing the C-GWAS results with the published GO-2 results. Across the defined 1-Mb regions surrounding these loci, we systematically evaluated 493 genes, of which 136 were supported by at least one line of evidence and 28 were prioritized as likely effector genes (**Supplementary Table 7**). Four genes (*TF (rs185357408), SLC2A1 (*rs61296119*), CREB3L2 (rs1646555),* and *CAPZB (rs10753560)*) showed the strongest credibility, with each supported by both variant-to-gene and gene-to-phenotype evidence. For example, for *SLC2A1 (*rs61296119), variant-to-gene evidence indicated that C-GWAS signals (lead variant: rs61296119) were predicted to affect *SLC2A1* expression by the ABC model, which predicts enhancer–gene links by combining regulatory activity with three-dimensional chromatin contact [22, 23] (**Supplementary Table 7**). This prediction was confirmed by the significant colocalization between C-GWAS signals and synovium methylation QTL of *SLC2A1* from osteoarthritic synovium. Concordantly, gene-to-phenotype evidence showed significantly different *SLC2A1* methylation levels in osteoarthritic chondrocytes compared to controls, and *SLC2A1* mutation is linked to human musculoskeletal phenotypes (**Supplementary text**). Meanwhile, *SLC2A1* (rs61296119) was associated with osteoarthritis at knee and thumb (**Supplementary Table 4**). *SLC2A1* encodes GLUT1, a glucose transporter abundantly expressed in chondrocytes[24]. Its inhibition in animal models has been shown to weaken chondrocyte function and induce joint degeneration[25]. Detailed evidence for *SLC2A1, TF, CREB3L2*, and *CAPZB* is described in **Supplementary Table 7**. *CREB3L2* (cAMP-responsive element-binding protein 3-like 2, rs1646555) plays an essential role in chondrocytes; supporting chondrocyte proliferation and extracellular matrix production, helping maintain cartilage homeostasis[26]. *CREB3L2* is located downstream of *SOX9*, a well-known osteoarthritis gene[27, 28]. *CAPZB* (rs1646555) encodes the β subunit of the heterodimeric CapZ complex that binds the barbed ends of actin filaments. In muscle cells, CapZ anchors actin at the Z-line and helps maintain properly formed, stable sarcomeres as muscles contract and relax[29]. This is relevant to osteoarthritis because muscle function is involved in osteoarthritis pathophysiology[30]. *TF* (rs185357408) encodes transferrin, a protein responsible for iron transport. Iron overload has been proposed as a contributor to osteoarthritis pathogenesis through oxidative stress and cartilage degeneration[31].

For the 46 novel signals located in 28 known loci, since they were all secondary signals in the known loci, we were first interested in whether they pointed to the same effector gene as the primary signals. By comparing effector gene prioritization results from GO-2, we found 12 genes were supported by both primary signals found in GO-2 and secondary signals found by this study: *CHST3 (*10:73721931_indel_G*), GDF5 (rs11905542), UQCC1(rs11905542), FAM53A(rs6599398, rs73188108, rs736436), GNG7 (rs55848295), JMJD4(rs12142953), NEK4 (rs11130328), PAPPA(rs803919), ALDH1A2* (rs563421104)*, NT5DC2(rs11130328), PBRM1(rs11130328),* and *PIK3R1 (rs11957631) (***Supplementary Table 8**). This observation has reinforced their candidacy as osteoarthritis effectors. For example, *ALDH1A2* (rs563421104) was among the highest-confidence OA genes in GO-2, supported by 11 lines of evidence types. Here, C-GWAS (rs563421104) signals were significantly colocalized with ALDH1A2 protein level at low-grade osteoarthritis cartilage pQTL (**Supplementary Table 8**). Meanwhile, C-GWAS also identified *ALDH1A2* (rs563421104) as a multi-trait signal, significantly associated with osteoarthritis at three of the five joints, namely knee, thumb and finger (**Supplementary Table 4**), further strengthening its role as an osteoarthritis gene.

In addition, these 46 novel secondary signals led to the prioritization of 29 likely effector genes that were not previously classified as high-confidence in GO-2 (**Supplementary Table 9**). For example, *BTN3A2* (rs2073531) had no supporting evidence from GO-2. Yet, the C-GWAS signal (rs2073531), which was associated with osteoarthritis at knee and hip (**Supplementary Table 4**), was predicted to affect *BTN3A2* gene expression by the ABC model, and colocalized with *BTN3A2* signals in a total of 6 different QTLs from osteoarthritis cartilage and synovium (**Supplementary Table 9**). Previous studies have identified *BTN3A2* as an immune-regulatory molecule that modulates γδ T-cell activation[32, 33], but its role in osteoarthritis was previously unknown. Finally, an additional nine novel effector genes were identified. They were not investigated in GO-2 due to differences in the locus range (**Supplementary Table 9**). In total, this annotation step linked the 66 novel signals to 66 likely effector genes.Notably, a subset of these newly prioritized genes also mapped to osteoarthritis-related biological processes highlighted by GO-2, including cartilage and chondrocyte biology, bone development, cellular signaling, and circadian regulation **(Supplementary Table 10)**, supporting that Trident can identify biologically plausible osteoarthritis-associated signals and genes.

Next, we examined the biology underlying the 19 replicated association signals with discordant multi-joint effects. Pathway enrichment analysis of the 40 likely effector genes linked to these 19 signals across 14 loci identified four significantly enriched biological processes (Supplementary Table 11), which clustered into three broader themes: immune receptor signaling, wound healing, and embryonic development. The two strongest enrichments were immune-related, both driven by *BTN3A2, BTN2A2, BTN1A1, BTN3A1, TRIM32*, and *TLR4*, whereas wound healing involved *NOTCH2, DSP, DGKI, TLR4*, and *ANGPTL2*, and embryonic organ development included *TBX15, NOTCH2, REST, ALDH1A2,* and *SOX9*. Together, these findings suggest that discordant effects across different osteoarthritis joints may arise from joint-specific differences in immune regulation, tissue repair, and developmental programs.

## Discussion

In this study, we addressed a practical challenge in multi-trait GWAS: how to evaluate the robustness of the obtained findings. We developed a three-step framework Trident that uses an earlier and smaller GWAS meta-analysis (**GO-1**) to test whether related phenotypes can be robustly combined, the latest and largest GWAS meta-analysis (**GO-2**) of the same phenotypes for discovery, and translational annotation to assess disease relevance and prioritize likely effector genes. We applied Trident to osteoarthritis at five joints and showed that it can support both robust multi-trait analysis and additional genetic discovery. In doing so, we showed that osteoarthritis genetic susceptibility is partly shared across joints, but not uniform, with some variants exerting opposite effects at different sites. Together, these findings establish Trident as a practical framework for robust multi-trait GWAS meta-analysis and illustrate its value for refining the genetic architecture of complex traits and diseases with consequence for future therapy.

Our results show that Trident can deliver robust multi-trait GWAS findings in the example of osteoarthritis. In the validation step, C-GWAS achieved a true-positive rate of 97.1%, while also recovering 81.2% of previously reported GO-1 signals at genome-wide significance. This indicates that C-GWAS performs robustly in large-scale osteoarthritis GWAS meta-analysis, increasing discovery while retaining established associations. When applied to the latest and largest osteoarthritis GWAS meta-analysis, Trident identified 66 novel signals. The credibility of these findings was supported by translational annotation. Although this step was designed primarily to prioritize effector genes, the evidence used was strongly osteoarthritis-relevant, including variant-to-gene links derived from osteoarthritis molecular data (colocalization with QTLs) and gene-to-phenotype links connected to osteoarthritis-related phenotypes. Thus, the fact that many of the identified signals converged on osteoarthritis-relevant genes and biological processes supports that Trident yields findings that are not only statistically robust, but also biologically coherent.

Applied to osteoarthritis, Trident contributed at two levels. First, it expanded the known genetic landscape of the disease by identifying 66 novel signals and prioritizing 66 likely effector genes. Second, it provided a more resolved view of osteoarthritis genetic architecture. Importantly, a significant C-GWAS association provides evidence that a variant is associated with at least one input phenotype, but does not by itself identify the joint(s) underlying the association. We therefore interpreted each C-GWAS signal alongside its association estimates from the five joint-specific GWAS. These analyses showed that 220 signals were associated with more than one joint, indicating that osteoarthritis at different joint sites shares part of its genetic basis. At the same time, 42 of these multi-joint signals showed discordant directions of effect across joints, suggesting that opposite genetic effects are not isolated observations but may represent a genuine feature of osteoarthritis biology. This distinction may inform the prioritization of future therapeutic studies: pathways acting consistently across joints may represent more promising targets for broadly effective osteoarthritis therapies, whereas pathways showing discordant effects across joints could be less suitable for general therapeutic targeting. Together, these findings show that multi-trait GWAS not only increases discovery in osteoarthritis but also reveals a biologically and potentially clinically relevant layer of cross-joint disease architecture relevant for future therapy.

Beyond osteoarthritis, multi-trait GWAS has the potential to become an important component of future consortium-scale GWAS meta-analysis. One clear advantage is increased discovery power. In our study, integrating information across related osteoarthritis phenotypes with C-GWAS achieved a 19.5% increase in discovery power compare to previous single trait GWAS meta-analysis[3], showing that additional discovery can still be achieved from existing data beyond gains from sample size alone. A second advantage is that multi-trait GWAS provides information on how variants, genes, and pathways act across related phenotypes, offering a more resolved view of disease biology, which is not available when analyzing each phenotype separately. Trident may therefore be useful also in other complex traits and diseases where related phenotypes share an underlying genetic basis. Cardiovascular disease represents one example, where coronary artery disease, stroke, heart failure and hypertension are clinically distinct, yet overlap in disease course, with one condition often predisposing to another[34]. With the success of GWAS meta-analyses of individual cardiovascular traits [35–38], building polygenic risk prediction score (PRS) is now an active area of the field. Yet, one main limitation of current cardiovascular PRS is that they are trait-specific, treating coronary artery disease[39], stroke[40], heart failure[41] and blood pressure[42] separately. As a result, prediction may be less complete because shared genetic effects across related outcomes are distributed across separate PRS, and less clinically useful because overall cardiovascular risk is assessed through multiple fragmented scores. By helping define this shared genetic basis, Trident could support the future development of unified cardiovascular PRS that provide more complete risk prediction and are thus more clinically useful.

Several limitations of our study should be considered. First, osteoarthritis phenotypes across contributing studies were defined using a mixture of criteria, which may have introduced phenotype heterogeneity into the analysis. Yet, C-GWAS identified 66 novel osteoarthritis associations. Second, most participants were of European ancestry, and the generalizability of these findings beyond Europeans remains to be established. Finally, although multiple lines of evidence supported the prioritized effector genes, these assignments remain probabilistic rather than causal, and experimental studies will be required to define their roles in osteoarthritis biology. Nevertheless, this evidence-based prioritization provides a rational shortlist of candidate genes for experimental studies aimed at establishing their roles in osteoarthritis biology.

In summary, we present a practical framework Trident for conducting and evaluating multi-trait GWAS meta-analysis that combines robustness assessment with enhanced genetic discovery, which is available for studying any complex trait and disease, particularly those with several biologically linked phenotypes. Applied to the example of osteoarthritis at five different joints, Trident identified novel risk signals and prioritized likely effector genes, while showing that osteoarthritis genetics is both shared and partly discordant across joints. Together, these findings provide a more resolved view of osteoarthritis genetic architecture and a practical foundation for future biological and translational studies.

## Data Availability

All data produced in the present study are available upon reasonable request to the authors

## Acknowledgments

Joyce van Meurs and Cindy G. Boer are supported by ReumaNederland (Project number: LLP-34). Yahong Wu and Ziyi Xiong were supported by the China Scholarship Council (CSC) fellowship for their Ph.D. studies at Erasmus MC University Medical Center Rotterdam. Benjamin Faber is supported by a Wellcome Trust Early Career Award (316390/Z/24/Z) and an Academy of Medical Sciences Starter Grant (SGL030\1057). Medical Ethics Committee of the Erasmus Medical Center has gave ethical approval for this work.

## Methods

### Data sources and phenotype selection

GWAS meta-analysis summary statistics were obtained from the Genetics of Osteoarthritis Consortium, which reported two large osteoarthritis GWAS meta-analyses, GO-1 and GO-2, as previously described[2, 3]. GO-1 included up to 826,690 individuals (177,517 cases and 649,173 controls) from 13 cohorts, predominantly of European ancestry. GO-2 included up to 1,962,069 individuals (489,975 cases and 1,472,094 controls) across 87 datasets from six ancestry groups, with most participants of European ancestry[2, 3]. Both studies analyzed 11 osteoarthritis phenotypes, including five joint-specific phenotypes—hip, knee, spine, finger and thumb osteoarthritis—and six additional phenotypes that partially or fully overlapped with these joint-specific traits: all osteoarthritis, total hip replacement, total knee replacement, knee and/or hip osteoarthritis, total joint replacement, and hand osteoarthritis[2, 3]. The C-GWAS analyses were based on the five joint-specific osteoarthritis GWAS meta-analysis summary statistics from GO-1 and GO-2: hip, knee, spine, finger and thumb osteoarthritis. These phenotypes were selected because they represent anatomically distinct manifestations of osteoarthritis and were expected to share part of their genetic basis. For C-GWAS input, variants were retained only if they were present in all five joint-specific summary statistics and were observed in at least 10% of the samples for each phenotype, consistent with previous applications of C-GWAS[5].

### C-GWAS analysis

C-GWAS was performed using the published R implementation of C-GWAS with default settings, as previously described[5]. Briefly, C-GWAS tests the null hypothesis that a variant has no effect on any of the input traits against the alternative hypothesis that it affects at least one trait. The method first estimates the correlation structure among the input GWAS summary statistics, then combines evidence across traits using a data-driven integration of two complementary association tests to generate a raw combined P value for each variant and finally calibrates these raw P values by simulation so that their null distribution is approximately uniform. Consequently, C-GWAS p-values are directly comparable to standard signal-trait GWAS, so the conventional genome-wide significance threshold of 5×10⁻⁸ was adopted as the study-wide significance threshold[6]. In this study, C-GWAS was applied separately to the GO-1 and GO-2 summary statistics for the five selected joint-specific osteoarthritis phenotypes.

### Defining independent signals and loci

Independent C-GWAS signals were identified using a two-step procedure consistent with the GO-2 study[3]. First, genome-wide significant variants were grouped by linkage disequilibrium (LD)-based clumping in PLINK2[43]using a P-value threshold of 5 × 10⁻⁸, a 2-Mb window, and an LD threshold of r² ≥ 0.1. LD was estimated using the UK Biobank v3 reference panel across all ancestries. Second, conditionally independent variants within each clump were identified using a customized P-value-based stepwise conditional analysis tool developed from COJO in GCTA[5, 44]. Two variants were considered independent if, after conditioning on the other variant, at least one remained genome-wide significant (P ≤ 5 × 10⁻⁸). Independent variants located within 1 Mb of each other were grouped into the same locus, and each locus was defined by extending 500 kb upstream and downstream of the outermost independent variants.

### Comparison of C-GWAS signals with reference GWAS catalogues

In both the validation and discovery analyses, independent signals identified by C-GWAS were compared with previously reported GWAS signals from GO-1 or GO-2 to determine whether they were already represented in the reference results or constituted **novel** independent associations. For each comparison, a C-GWAS locus was first classified as known if it contained at least one reference signal; otherwise, it was classified as novel. C-GWAS signals located in novel loci were classified as novel signals. Within known loci, pairwise LD was calculated between each C-GWAS signal and the reference signals. A C-GWAS signal was considered to represent a known signal if it was in high LD (r² ≥ 0.75) with at least one reference variant. For C-GWAS signals not in high LD with any reference signal in the same locus, conditional analysis was performed using the same customised P-value-based software described above[5, 44]. A C-GWAS signal was considered novel if it was independent from all reference signals in that locus, and a reference signal was considered represented if it was not independent from at least one C-GWAS signal. If a C-GWAS signal was unavailable for conditional analysis, a proxy variant in high LD (r² ≥ 0.8) was used. Based on this procedure, C-GWAS signals were classified into three categories: novel signals in novel loci, novel signals in known loci, and known signals in known loci.

### True-positive assessment

We assessed the true-positive rate of C-GWAS resulting from GO-1. A C-GWAS signal was considered true positive if it was represented by either GO-1 or GO-2. First, the 70 independent C-GWAS signals identified in GO-1 were compared with the published GO-1 signal catalogue using the comparison framework described above. This identified 59 signals that were already represented in at least one of the 11 osteoarthritis phenotype GWAS meta-analyses in GO-1. The remaining 11 GO-1-novel C-GWAS signals were then evaluated in GO-2[3] by looking up their association results across all 11 osteoarthritis GWAS meta-analyses. For each signal, the minimum P value across the 11 GO-2 phenotypes was recorded. A signal was considered represented by GO-2 if its minimum P value was ≤ 4.13 × 10⁻⁴, corresponding to a Bonferroni-corrected threshold of 0.05 divided by 11 signals and 11 phenotype lookups.

### False-negative assessment

To assess the false-negative rate of C-GWAS resulting from GO-1, previously reported independent GO-1 osteoarthritis signals were used as the reference set. Signals that were excluded from the C-GWAS input during variant filtering and harmonization were removed before analysis. The remaining GO-1 lead signals were then looked up directly in the full GO-1 C-GWAS summary results, and recovery was assessed at two thresholds: genome-wide significance (P < 5 × 10⁻⁸) and suggestive significance (P < 1 × 10⁻⁶).

### Discovery analysis

To identify novel osteoarthritis signals of C-GWAS resulting from GO-2, independent C-GWAS signals resulting from GO-2 were compared with the complete GO-2 signal catalogue across all 11 osteoarthritis phenotypes using the comparison framework described above. Signals classified as novel signals in either novel or known loci were considered novel osteoarthritis associations.

### Benchmark analysis

To quantify the discovery gain provided by C-GWAS, the GO-2 C-GWAS signals were compared with two additional GO-2 reference catalogues: (i) the signal set identified from the same five joint-specific osteoarthritis phenotypes used as C-GWAS input, and (ii) the signal set identified from the single “All osteoarthritis” GWAS meta-analysis. These comparisons were used to estimate the number of additional signals identified by C-GWAS relative to conventional single-trait GWAS analyses approaches.

### Classification of multi-trait and discordant signals

To characterize the joint-specific association patterns underlying the GO-2 C-GWAS results, each of the 297 independent genome-wide significant C-GWAS lead variants was evaluated in the five GO-2 joint-specific osteoarthritis GWAS meta-analysis summary statistics: hip, knee, spine, finger and thumb osteoarthritis. Alleles and effect directions were harmonized across phenotypes using the GO-2 per-trait summary statistics. For each lead variant, association with each joint-specific phenotype was assessed using a Bonferroni-corrected nominal significance threshold of P ≤ 0.01 (0.05 ÷ 5 phenotypes). A C-GWAS signal was defined as a multi-trait signal if it was significantly associated with at least two joint-specific osteoarthritis phenotypes. Among multi-trait signals, direction of effect was evaluated by comparing the signs of the beta coefficients across significant phenotypes. Signals showing the same effect direction across associated phenotypes were classified as directionally consistent, whereas signals showing opposite effect directions across at least two associated phenotypes were classified as directionally discordant.

### Sensitivity analysis

To assess whether discordant multi-joint effects were robust, we performed a sensitivity analysis using osteoarthritis GWAS summary statistics from the Rotterdam Study, in which osteoarthritis was defined radiographically. This analysis included the three Rotterdam Study sub-cohorts that also contributed to GO-2[3]. Summary statistics from these three sub-cohorts were first meta-analysed using the same approach as applied in GO-2[3]. The 42 directionally discordant GO-2 C-GWAS signals were then evaluated in the Rotterdam Study meta-analysis. For each signal, the lead variant was looked-up after harmonization to the same effect allele as in GO-2; if the lead variant was unavailable, a proxy variant in high LD (r² ≥ 0.8) was used. Support for discordance was assessed qualitatively by comparing the cross-joint direction-of-effect pattern observed in GO-2 with that observed in the Rotterdam Study meta-analysis. A signal was considered supported if the same discordant pattern of effects across joints was observed in the Rotterdam Study, irrespective of statistical significance.

### Prioritization of likely effector genes

To assess the biological relevance of the detected novel osteoarthritis associations and prioritize their likely effector genes, we integrated 22 lines of translational evidence using the same general pipeline as GO-2[3]. These lines of evidence were grouped into two broad categories: variant-to-gene evidence and gene-to-phenotype evidence. Variant-to-gene evidence captured links between association signals and candidate target genes, including Activity-By-Contact (ABC) model predictions and colocalization with molecular quantitative trait loci in osteoarthritis-relevant tissues. Gene-to-phenotype evidence captured support for the involvement of candidate genes in osteoarthritis-relevant biology and related phenotypes in humans and mice. Full details of all datasets, evidence lines and analytical thresholds are provided in the Supplementary Methods. This translational evidence framework was applied to all genes within the 20 novel loci and the 28 known loci. For each candidate gene, support was summarized across the 22 evidence lines. A gene was prioritized as a likely osteoarthritis effector gene if it was supported by at least one line of variant-to-gene evidence or by three or more lines of gene-to-phenotype evidence. For loci already reported in GO-2[3], the likely effector genes prioritized from the C-GWAS secondary signals were additionally compared with the corresponding effector-gene prioritization results from GO-2 to determine whether the same genes were supported by both analyses.

### Pathway analysis

To place the newly prioritized osteoarthritis genes in biological context, we first assessed whether the 66 likely effector genes linked to the 66 novel C-GWAS signals mapped to osteoarthritis-related biological processes previously highlighted by GO-2[3]. In addition, to investigate the biology underlying discordant multi-joint effects, we performed pathway enrichment analysis on the likely effector genes linked to the 19 discordant signals that showed the same cross-joint direction-of-effect pattern in the Rotterdam Study sensitivity analysis. Gene Ontology Biological Process enrichment was performed using the clusterProfiler package in R[45].

